# The development of a national bilingual cross-sectional questionnaire on attitudes towards supervised consumption sites and e-health overdose response interventions in Canada: The Canadian National Questionnaire on Overdose Monitoring (CNQOM)

**DOI:** 10.1101/2025.03.27.25324132

**Authors:** N. Rider, B. Seo, D. Viste, W. Rioux, N. Sedaghat, B. Pan, Y.N. Al Hamarneh, G.R. McCormack, F. Aghajafari, L. McDougall, S. M. Ghosh

**Affiliations:** Faculty of Medicine, Cumming School of Medicine, University of Calgary, 3330 Hospital Dr NW, Calgary, AB T2N 4N1, Canada; Department of Medicine, Faculty of Medicine & Dentistry, University of Alberta, 2J2.00 Walter C Mackenzie Health Sciences Centre, 8440 112 St. NW, Edmonton, AB T6G 2R7, Canada; Epidemiology Coordinating and Research (EPICORE), Department of Medicine, Faculty of Medicine & Dentistry, University of Alberta, 2J2.00 Walter C Mackenzie Health Sciences Centre, 8440 112 St. NW, Edmonton, AB T6G 2R7, Canada; Department of Pharmacology, Faculty of Medicine and Dentistry, University of Alberta, 362 HMRC Building, Edmonton, AB T6G 2S2, Canada; Department of Community Health Sciences, Cumming School of Medicine, University of Calgary, 3330 Hospital Dr NW, Calgary, AB T2N 4N1, Canada

## Abstract

**Background:** Supervised consumption sites (SCSs) and overdose response hotlines/applications (ORHAs) are harm reduction interventions aimed at reducing fatal overdose mortality. Little is known about key informant perspectives regarding these services. Herein, our objective is to describe the process of developing, testing, and distributing a novel national online questionnaire to measure perspectives and attitudes towards SCS and ORHAs among key informants in Canada.

**Methods:** A bilingual online instrument (the *Canadian National Questionnaire on Overdose Monitoring,* CNQOM) was developed, pilot tested and revised with the aim of ensuring content and construct validity. Questionnaire respondents were recruited nationally from four key informant groups (People Who Use Substances, health professionals, emergency responders, and the general public) using a mix of purposive and representative sampling strategies. Respondents came from every province and territory in Canada, and respondents from the general public were proportionally represented. A stepwise response validation approach was used to remove invalid questionnaire responses. Test-retest reliability of instrument questions was assessed using Spearman’s Rank Correlation, the Wilcoxon Rank Sum Test, and Cohen’s Kappa.

**Results:** A total of 4,445 valid responses was obtained from the four key informant groups following data cleaning. Test-retest reliability of instrument questions demonstrated slight to substantial stability in responses.

**Discussion:** The CNQOM is the first online questionnaire in Canada designed to capture perspectives and attitudes towards SCSs and ORHAs among diverse key informant groups. Our questionnaire was administered to a large, geographically diverse sample and designed to capture the perspectives of four key informant groups. Lower than expected test-retest reliability may be explained by lack of participant familiarity with SCS and especially ORHAs and the impersonal nature of the instrument content among some respondents. Future work will elucidate key informant perspectives on these services based on the data.

## BACKGROUND

Fatal drug poisonings, also known as drug overdoses, are responsible for an ongoing public health crisis in Canada (1). In both Canada and the United States, the majority of opioid-related deaths result when the victim is using substances alone, a major risk factor for fatal overdose (2,3). Stigma, multidrug use, and use in an unfamiliar environment have been associated with using substances alone (4), and relatively few injection drug users - only 26% in one study - never used alone (5). During the COVID-19 pandemic, People Who Use Substances (PWUS) may have experienced more pressure to use alone due to fear of disease transmission or exposure, social and physical isolation, and recommended quarantine and isolation measures (6). Partially in response to these pressures, many providers of novel overdose detection technologies (ODTs) began to expand their services in North America (7). ODTs are e-health or virtual harm reduction technologies that include: fixed-location devices such as sensors, motion detectors, intercom systems, and buttons; mobile phone applications; hotlines; and wearable technologies such as temperature sensors, movement detectors, and automatic antidote (naloxone) injectors (8). At this time, there is limited evidence regarding the safety, effectiveness, costs, and practicality of these technologies (7,8). Overdose response hotlines and applications (ORHAs) are subcategories of ODTs that share a number of similarities. Both overdose response hotlines (ORHs) and overdose response applications (ORAs) allow individuals to be monitored remotely while they use substances and can activate an emergency or community response in the event of an overdose (7).

### Rationale

Proctor’s Framework is a tool designed to aid in the assessment of implementation outcomes based on various key evaluation categories such as acceptability, adoption, appropriateness, costs, feasibility, fidelity, penetration, and sustainability (9). Within Proctor’s Framework, acceptability is “the perception among implementation stakeholders that a given treatment, service, practice, or innovation is agreeable, palatable, or satisfactory” (9). The use of an implementation science approach to new technologies can be useful in identifying areas requiring further study or quality improvement. Consequently, understanding the acceptability of overdose response hotlines and applications will contribute to their evaluation.

Overall, there is a limited amount of research detailing the acceptability of ORHAs to key informants. Hotlines and applications have been studied only in a limited fashion, while fixed-location devices have received even less attention from an acceptability perspective (8,10,11). Most research on ORHA acceptability has been conducted through qualitative interviews or small-scale questionnaires about specific interventions; furthermore, such work has largely focused on PWUS and healthcare providers (10,12–14). There have not been any large-scale quantitative studies of ORHA acceptability amongst key informants, or an assessment of the perspectives of first responders and the general public.

In contrast to the lack of information about the acceptability of ORHAs, attitudes towards supervised injection facilities/supervised consumption sites (SIFs/SCS) are better established (15). SIFs/SCS are a well-established harm reduction intervention that is demonstrated to be safe and effective in preventing drug poisoning fatalities, bloodborne pathogen transmission, and improving access to community programming (16–19). In their systematic review of qualitative studies, Lange and Bach-Mortensen (2017) found that the acceptability of SCS had been assessed amongst a wide variety of key partners - most commonly, people who use substances (PWUS; 37 studies), first responders (22 studies), health professionals (15 studies), substance use sector employees (10 studies), and SIF employees (9 studies). Other groups (e.g. business sector, government officials) were less frequently studied. Of the 47 studies included in Lange and Bach-Mortensen’s review, only 33 were peer-reviewed, with the remainder consisting of equal numbers of reports and dissertations. These findings support PWUS, first responders, and health professionals as key stakeholders in supervised consumption. A recent survey of Canadian adults found the majority supported the provision of a variety of harm reduction interventions, including outreach, naloxone provision, drug checking services, sanitary needle distribution, and supervised drug consumption; however, there was notably less support for opioid agonist treatment without abstinence (low-threshold agonist therapy) and safe inhalation (20).

Public and political acceptability have important implications for the successful adoption of public health interventions as diverse as sugar-sweetened beverage taxation, policies targeting physical activity, malaria drug administration programs, and vaginal ring use (21–24). Assessing the acceptability of SCS and ORHAs will be critical when considering whether future investment and ongoing funding are politically palatable. Furthermore, knowledge of attitudes towards established harm reduction interventions in the SCS literature can help guide a quantitative approach to assessing attitudes towards ORHAs while simultaneously providing a baseline against which perceptions towards ORHAs can be compared.

### Research Aims

The methodology described herein is part of a diverse and comprehensive assessment of implementation of ORHAs by our team. We have named our questionnaire the Canadian National Questionnaire on Overdose Monitoring (CNQOM). The CNQOM will be used to gather information on the perspectives and attitudes towards ORHAs and SCS among key informants from the literature (PWUS, first responders, health care providers, and the general public). Informing the development of the CNQOM was our aim to create a valid instrument to facilitate the:

1. Assessment of public awareness of ORHAs and SCS.
2. Evaluation of attitudes of key informants regarding ORHAs and SCS and their appropriateness.
3. Comparison of attitudes of key informants towards ORHAs to attitudes towards SCS.
4. Assessment of the preferences of key informants regarding ORHAs and SCS.
5. Identification of elements that key informants believe may be essential for safety, client satisfaction, acceptability, and quality improvement of ORHAs.
6. Assessment of barriers to using ORHAs and ways to increase awareness about ORHAs.
7. Assessment of respondent experiences with ORHAs available in Canada.

In the current paper, we describe the methodology used to develop, distribute, validate, and assess the reliability of the CNQOM.

## METHODS

### Study Questionnaire

#### Development

The CNQOM development approach followed steps 1-5 proposed for validation of a recent nutrition knowledge instrument (25). Purification and refinement activities (step 6) were not conducted since there was no “correct” answer to the questions in the CNSORHA, and the questionnaire measures clearly discrete constructs. Similarly, assessment of factorial structure and dimensionality (steps 7a and 7b) was not performed. Temporal stability (step 8) was assessed through a test-retest reliability analysis (see below). A literature search did not yield any previous instruments suitable for the scope of the current project. Similarly, no appropriate instrument existed to measure in-depth attitudes towards SCS. A new study-specific instrument was developed by the study team and informed by key partners including line operators and senior executives associated with Canada’s National Overdose Response Service (NORS - an overdose response hotline), an addictions medicine physician (MG), harm reduction personnel, people with lived experience of substance use, first responders, health care providers, and members of the general public.

Each question was developed to measure a unique aspect of perceptions regarding SCS and/or ORHAs. Question development was informed by elements of Proctor’s Framework for Implementation Science (9), a systematic review of attitudes towards SIFs (15), and the opinions of key partners and the study team based on their expert knowledge of ORHAs and SCSs. Furthermore, questions were informed by preliminary qualitative data from a series of studies on ORHAs that were in progress by the same research team to permit cross-method consistency. To enable a direct comparison of attitudes between SCS and ORHAs, the questionnaire contained identically worded questions assessing perceptions of key elements of both interventions. Once the instrument was finalized, a professional translation service was used to translate the instrument from English to French (both of which are official languages in Canada) to allow bilingual administration.

#### Instrument Components and Questions

The online questionnaire included seven sections (450 questions total; see below): (1) front matter and consent (8 questions); (2) general demographics (14 questions); (3) substance use and mental health demographics (for participants identified as PWUS only;15 questions); (4) questions comparing SCS to ORHAs (96 questions); (5) overall intervention acceptability for SCS and ORHAs (16 questions); and (6) questions pertaining to specific elements of ORHAs (most of which were only asked to participants identifying as PWUS, first responders, or healthcare providers; 77 questions). The seventh and final section of the instrument asked questions about specific ORHAs to individuals with experience using them (221 questions about 5 different services). Questions about specific services were solely asked to individuals who had used those services. There were two trap questions to assess respondent attention. At the end of the questionnaire, there was an open text box asking for any additional comments. Twenty-four metadata elements (e.g. start time, time to complete) were generated by Qualtrics XM (an online survey platform). Participants had the option to provide their email, indicating their willingness to be contacted for future research. Response options for instrument questions included multiple-choice, choose all that apply, balanced Likert scale, and numeric formats (Rattray & Jones, 2007). The questionnaire was designed to be approximately 10-15 minutes in length based on initial testing conducted by the study team and Qualtrics derived metrics. For PWUS, healthcare providers, and the first responders, the survey was estimated to take slightly longer (15-25 minutes) due to additional questions aimed at these groups. Due to the novelty of the services studied, most questions included a “not sure” or neutral option.

#### Pilot Testing

Three rounds of pilot testing were conducted by the principal investigator (MG) and the study team, which included a resident physician (NR), post-doctoral fellow (TM), research assistants (DV, WR, BS), healthcare professionals, first responders, members of the general public, and harm reduction workers/people with lived experience of substance use to ensure the use of appropriate and non-stigmatizing language, thoroughness of content, and readability. Feedback was incorporated into the questionnaire during each round of pilot testing.

#### Instrument Validity

The face validity of each question was verified by representatives from the four key partner groups to ensure proper construct conceptualization (26). Content and construct validity was enhanced *a priori* by experts in the field of harm reduction and addiction medicine (27). Key partners and experts were consulted on each question and asked to provide feedback to improve question discrimination. As no similar measures were available, it was not possible to assess convergent validity.

### Participants

Eligible participants had to reside in Canada, be at least 18 years of age, and be able to read and respond in English or French; participants who did not meet these criteria were excluded. Furthermore, participants had to belong to one of four key informant groups - PWUS (those with lived experience of personal substance use), health professionals, emergency responders, or the general public. While not specifically targeted, some participants had adjacent harm reduction or substance use experiences including being family members of people with substance use concerns, working as social workers with vulnerable populations, and working in justice departments or with government agencies that directly interact with substance-using populations. A $15 CAD honorarium was given to PWUS (those with lived or living experience of substance use), who completed the questionnaire using links from specific partner organizations through e-transfer or gift card (in accordance with partner organization policy). Only versions of the questionnaire intended for distribution to PWUS were eligible for payment to avoid “gaming” of the payment system. Sampling was undertaken using a combination of purposive and random approaches to ensure sufficient representation from all four key informant groups. To facilitate access to potential participants from each of these groups, the study team compiled a list of 610 potential partner organizations that might have contacts in the first three key informant groups (Table 1). A standardized email was sent to each of these organizations over a 13-month period starting on July 1^st^, 2022 and ending in May 31^st^, 2023. Organizations that responded to the initial email were sent additional information about the study. The study team also worked with organizations to complete any necessary paperwork to get the questionnaire distributed to the membership, employees, clients, or other relevant contacts of the partner organization. Partner organizations distributed the questionnaire by email or posters using a form letter or poster supplied by the research team and were responsible for promoting the questionnaire through internal channels. Distribution materials provided by the research team included a link to the questionnaire and a QR code, along with posters and email templates. For distribution to the final key informant group (general public), a professional marketing and research analytics company was contracted to obtain a large random Canadian sample with proportional representation from all provinces and territories. This company also assisted in inadvertently providing a limited number of healthcare professional, first responder, and PWUS respondents who were members of their pool (these were recategorized to the more appropriate stakeholder group). Furthermore, the company collected data for the main analysis as well as data for the purpose of conducting test-retest reliability analysis. The company offers nominal compensation (<$4 per person) in the form of points to their participant pool. The difference in compensation between the different groups reflects practical, funding, and equity considerations. Participants were offered the option to request their data to be withdrawn from analysis (participants who did not provide an email did not have this option due to response anonymity); however, no participants exercised this option.

**Table 1.**
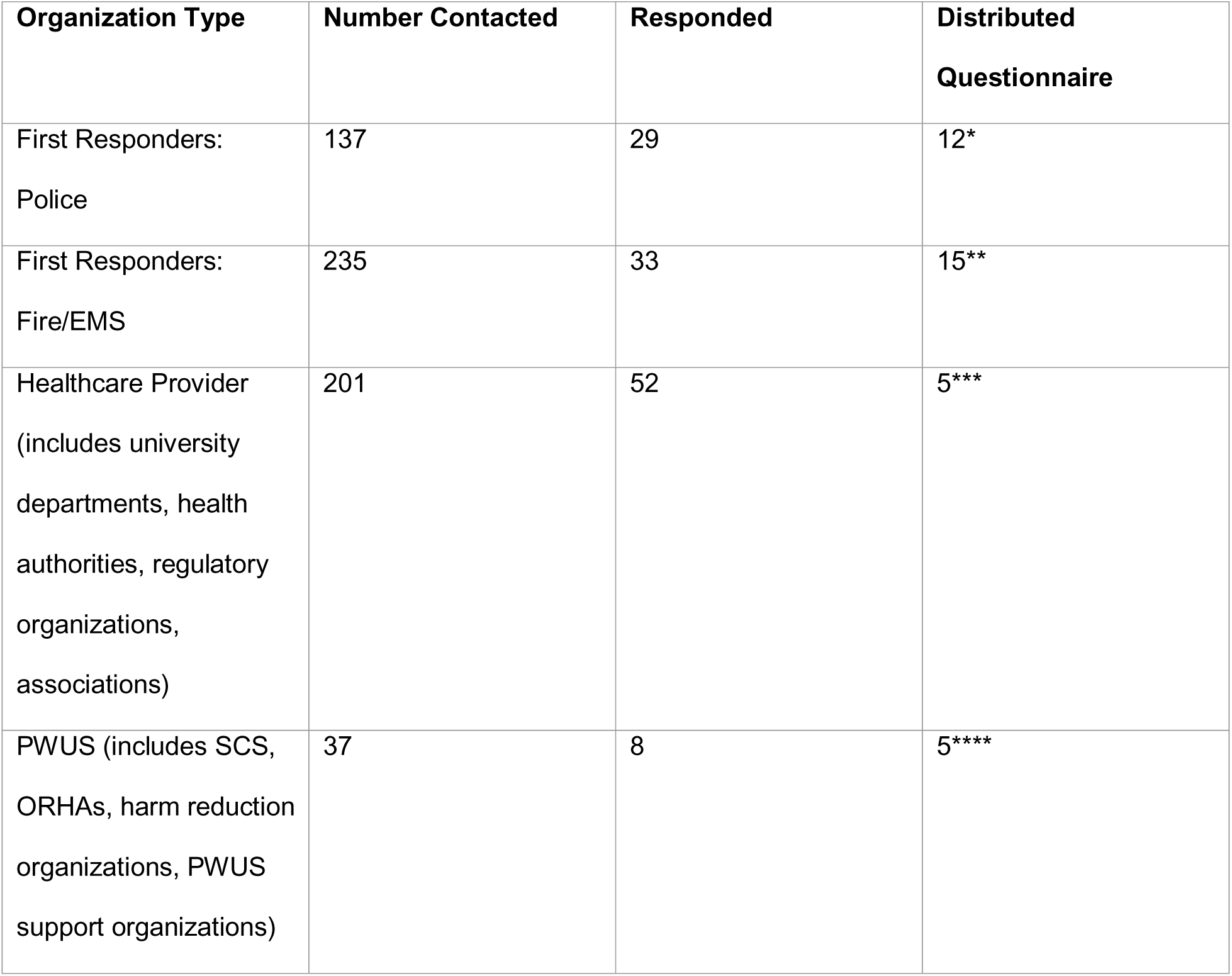
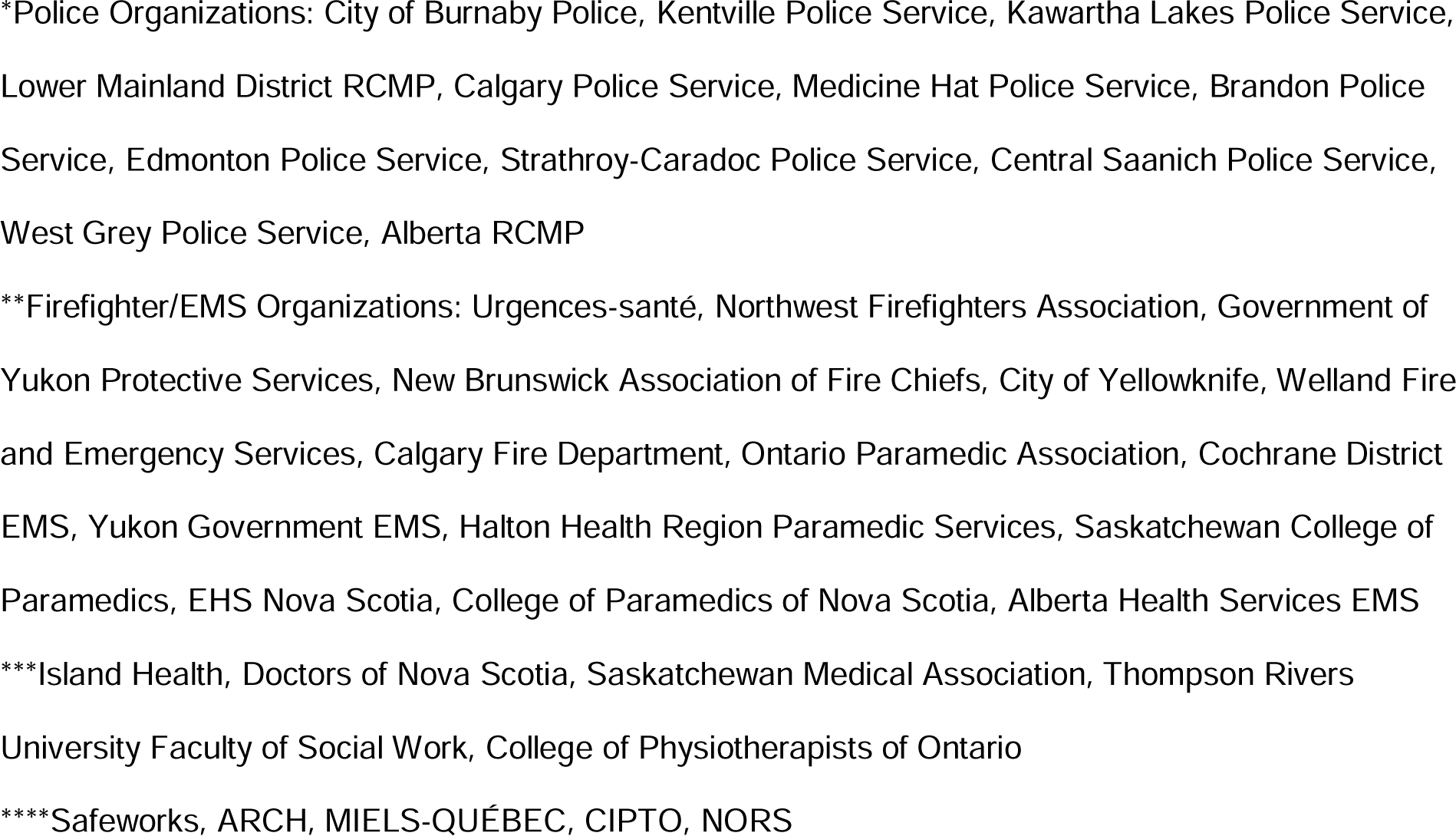
Organizations contacted by type, number of organizations that responded, and number of organizations that distributed the questionnaire.

| Organization Type | Number Contacted | Responded | Distributed Questionnaire |
| --- | --- | --- | --- |
| First Responders:<br>Police | 137 | 29 | 12* |
| First Responders:<br>Fire/EMS | 235 | 33 | 15** |
| Healthcare Provider<br>(includes university departments, health authorities, regulatory organizations, associations) | 201 | 52 | 5*** |
| PWUS (includes SCS, ORHAs, harm reduction organizations, PWUS support organizations) | 37 | 8 | 5**** |

\*Police Organizations: City of Burnaby Police, Kentville Police Service, Kawartha Lakes Police Service, Lower Mainland District RCMP, Calgary Police Service, Medicine Hat Police Service, Brandon Police Service, Edmonton Police Service, Strathroy-Caradoc Police Service, Central Saanich Police Service, West Grey Police Service, Alberta RCMP
\*\*Firefighter/EMS Organizations: Urgences-santé, Northwest Firefighters Association, Government of Yukon Protective Services, New Brunswick Association of Fire Chiefs, City of Yellowknife, Welland Fire and Emergency Services, Calgary Fire Department, Ontario Paramedic Association, Cochrane District EMS, Yukon Government EMS, Halton Health Region Paramedic Services, Saskatchewan College of Paramedics, EHS Nova Scotia, College of Paramedics of Nova Scotia, Alberta Health Services EMS
\*\*\*Island Health, Doctors of Nova Scotia, Saskatchewan Medical Association, Thompson Rivers University Faculty of Social Work, College of Physiotherapists of Ontario
\*\*\*\*Safeworks, ARCH, MIELS-QUÉBEC, CIPTO, NORS

#### Sample Size

The total sample size for this study was calculated by summing all the required sample sizes for each of the study groups. For the general public, the required sample size using the population size of Canada (37,238,396) and the following assumptions 95% confidence interval and 5% margin of error was 601 (28). For the first responders, the required sample size using the population size of first responders in Canada and the same assumptions as above was 384 (29). Similar approach was used for PWUS and health care providers and the required sample sizes were 384 (29) and 385 (30) respectively, for a grand total of 1,754. It was ultimately necessary to collect excess responses from the members of the general public to achieve an adequate number of quality responses from the other key groups, resulting in a larger number of responses from the general public relative to other groups.

### Instrument Delivery and Security

The questionnaire was administered online using the Qualtrics XM platform to improve ease of distribution and affordability. It was optimized for both mobile and web viewing. Due to the provision of an honorarium to PWUS answering select versions of the questionnaire delivered by partner agencies known to have contacts who met the criteria for being PWUS, the research team added additional security measures to the questionnaire to prevent financial fraud, including ReCAPTCHA V3. Preliminary distribution resulted in an apparent bot attack on one version of the questionnaire distributed in September 2022, evidenced by a massive number of responses (about 4000) over only a few days which was deemed to be suspicious, given the poor response quality, a high number of attempted duplicate submissions, dubious looking emails (as assessed by the research team and email verification service Verifelia), and poor ReCAPTCHA V3 scores. Following this incident, additional security measures were added in consultation with the Qualtrics XM team, the University of Calgary’s Office of Institutional Analysis, and ethics. These measures included the addition of ReCAPTCHA V1 and V2 to the questionnaire, the enabling of IP address collection and duplicate response blocking foreign IP blocking, and the creation of even more versions of the questionnaire to enable easier detection of fraudulent responses. No e-transfers were offered for suspected fraudulent responses and the same was communicated to affected partner organizations. A similar incident occurred in January 2023. Data suspected to be fraudulent was stored for future reference, but not included in analyses. Once data collection was finished, all versions of the questionnaire were closed to further responses.

### Statistical Analysis

#### Response Validation

Validation activities were undertaken to ensure that all responses from both the main dataset and the test-retest reliability dataset were legitimate (Table 2). Respondent submissions which were considered potentially fraudulent (e.g. suspected duplicates) were removed using a stepwise approach. Responses underwent an initial screen using ReCAPTCHA data; respondents that failed ReCAPTCHA V1 or V2 were prevented from completing the questionnaire after the first page and these incomplete responses were removed. ReCAPTCHA V3 responses are scored from 0.0 to 1.0, with 0.0 almost certainly representing a software attempt and 1.0 being almost certainly human (Google, 2024). Though the default ReCAPTCHA V3 cutoff is typically 0.5, we used a more conservative cutoff of 0.9, which has previously been considered to be on par with human performance (Akrout et al., 2019) and was recommended by the survey distribution company.

**Table 2.** Results of response validation procedures demonstrating total questionnaire attempts and reasons for response removal.

|  |  |
| --- | --- |
| **Total attempts** | **9863** |
| Recaptcha score <0.9 | 507 |
| <5% progress on questionnaire | 842 |
| Under 18 years | 59 |
| IP from outside Canada | 9 |
| Proportional provincial quota already met | 3222 |
| Impossibly fast (<500 seconds) | 3986 |
| Duplicate respondents | 60 |
| **Total removed:** | **-4674** |
| **Total reaching 33%** | **5189** |
| Failed Trap question 1 | 84 |
| Failed Trap question 2 | 164 |
| **Total removed:** | **-248** |
| **Total reaching 41%** | **4941** |
| **Didn't reach 100%** | **-536** |
| **Total reaching 100%** | **4405** |
| **Trap questions waived upon examination of written response** | **+40** |
| **Grand Total** | **4445** |

Further responses were removed iteratively for not meeting inclusion criteria (assessed based on a series of questions on the first page of the questionnaire), having <5% progress in the questionnaire, having an IP address originating outside of Canada, completing the questionnaire in <500 seconds, being flagged by Qualtrics as duplicate responses through browser cookies, and failing one or both of two trap questions. Responses that were completed in <500 seconds were deemed likely fraudulent due to testing that showed respondents would struggle to thoughtfully respond to questions before the 41% completion mark if they completed the questionnaire faster than this. Subsequently, responses not reaching the 41% completion mark (representing the end of the fourth questionnaire section, which compared attitudes towards SCS and ORHAs) were also removed. A limited number of responses (40) were manually screened back into the sample due to clearly legitimate comments in the open text box at the end of the questionnaire, despite having failed one or both trap questions.

#### Data Coding and Descriptive Statistics

A data dictionary was created and the data were cleaned. The cleaning procedure included the removal of all identifying information (email addresses, any identifying free text responses). Statistical analysis was performed based on the as-observed principle and by using R 3.4.0 (Vienna, Austria; https://www.R-project.org/) and SAS 9.4 software (SAS Institute Inc. Cary, NC, USA). Descriptive statistics were calculated by the Alberta Strategy for Patient-Oriented Research Support Unit (AbSPORU). Missing data underwent pairwise deletion. A descriptive summary of demographic information was prepared.

#### Instrument Reliability Assessment

Test-retest reliability participants (n = 140) were recruited as a separate cohort from the same pool as the general public cohort and using the same eligibility criteria. Assuming a total sample of 140 individuals, we would have more than 90% power to detect a correlation coefficient of 0.4. The power calculation is calculated by correlation test using, Z transformation of the correlation coefficient, with the R package “pwr: Basic Functions for Power Analysis”. Since not all questions were deemed relevant for all groups of respondents, only questions asked to the general public were included in the reliability assessment. Missing data and “not sure” responses underwent pairwise deletion. Participants were sent the questionnaire twice and responses that were collected from a single participant 7-10 days apart were included in the reliability analysis.

Due to the ordinal nature of the questions, test-retest reliability was assessed using Spearman’s Rank Correlation, the Wilcoxon Rank Sum Test (α=0.05), and Cohen’s Kappa (31,32). Historically, Kappa (κ) values have been interpreted as follows in regard to the level of agreement: 0.00 - 0.20 (slight), 0.21 - 0.40 (fair), 0.41 - 0.60 (moderate), 0.61 - 0.80 (substantial), and 0.81 - 1.00 (almost perfect) (33), though cutoffs on the higher end of the scale may be preferred in some areas of research (McHugh, 2012). Correlation values (*r_s_*) of 0.00 to 0.30 are deemed negligible, 0.31 - 0.50 low, 0.50 - 0.70 moderate, 0.71 - 0.90 high, and 0.90 - 1.00 very high (34).

### Ethics and Reporting

Participation was voluntary and consent was obtained by electronic means. Participants read over a digital consent form and could select “I agree to participate” or “I DO NOT agree to participate”. The latter response ended the questionnaire. The study was approved by the University of Calgary’s Conjoint Health Research Ethics Board (CHREB; REB #21-1646). The STROBE Statement for cross-sectional studies was used in the drafting of this manuscript.

## RESULTS

### Response Validation

Out of 610 partner organizations contacted initially, 122 responded, but only 37 distributed the questionnaire (Table 1). A total of 9,863 participants attempted to complete the CNQOM. Of these 5,418 were excluded during the validation stage due to completing the questionnaire too fast to provide thoughtful responses, progressing <41% through the questionnaire, and having a low ReCAPTCHA score (indicating suspicious responses), resulting in 4,445 deemed to be of sufficient quality for a valid analysis (Table 2). While 536 responses of the total 4,445 validated responses did not reach 100% completion, it was determined that they were still of sufficient quality to be included in current and future analyses. After response validation, a broad main sample was obtained using a combination of purposive and a provincial representative approach (Table 3). A total of 749 participants completed the baseline test-retest questionnaire, and 460 completed the follow-up questionnaire between 7 and 15 days after the baseline questionnaire (mean = 9.08 days, median = 8.21 days). Test-retest respondents were not part of the main dataset. Initial response validation removed incomplete questionnaires, questionnaires completed from an IP address outside of Canada, and duplicate responses, leaving 336 baseline and 223 follow-up questionnaire responses. A unique identifier was then used to pair the baseline and follow-up questionnaires, yielding 140 completed participant paired questionnaires, for which demographic information is summarized (Table 4).

**Table 3:** Demographics of participants in the main dataset. Unless otherwise specified, numbers are counts with percentage of total respondents in brackets.

|  | General public | Emergency<br>service<br>providers | Health care<br>providers | People who<br>use<br>substances | Total |
| --- | --- | --- | --- | --- | --- |
| **Total** | 2866 | 372 | 381 | 826 | 4445 |
| **Age** |  |  |  |  |  |
| Mean | 48.8 | 40.2 | 38.9 | 39.7 | 45.5 |
| Standard deviation | 16.9 | 10.8 | 13.5 | 13.9 | 16.3 |
| **Gender** |  |  |  |  |  |
| Male | 1248 (43.5) | 236 (63.4) | 82 (21.5) | 373 (45.1) | 1939 (43.6) |
| Female | 1507 (52.5) | 131 (35.2) | 286 (75.0) | 401 (48.5) | 2325 (52.3) |
| Gender-diverse | 26 (0.9) | 2 (0.5) | 8 (2.0) | 49 (5.9) | 85 (1.9) |
| Transgender Male | 6 (0.2) | 0 (0) | 3 (0.7) | 13 (1.5) | 22 (0.4) |
| Transgender Female | 1 (0.0) | 1 (0.2) | 0 (0) | 5 (0.6) | 7 (0.1) |
| Non-binary | 18 (0.6) | 1 (0.2) | 5 (1.3) | 28 (3.3) | 52 (1.1) |
| Gender Queer | 0 (0) | 0 (0) | 0 (0) | 1 (0.1) | 1 (0.0) |
| Gender fluid | 1 (0.0) | 0 (0) | 0 (0) | 1 (0.1) | 2 (0.0) |
| Two-Spirit | 0 (0) | 0 (0) | 0 (0) | 1 (0.1) | 1 (0.0) |
| No Gender | 85 (2.9) | 3 (0.8) | 5 (1.3) | 3 (0.3) | 96 (2.1) |
| **Ethnicity** |  |  |  |  |  |
| White | 2115 (73.7) | 310 (83.3) | 269 (70.6) | 592 (71.6) | 3286 (73.9) |
| Indigenous | 99 (3.45) | 16 (4.30) | 22 (5.77) | 125 (15.1) | 262 (5.89) |
| Other Ethnicities | 524 (18.2) | 30 (8.06) | 81 (21.2) | 99 (11.9) | 669 (15.0) |
| Missing | 128 (4.4) | 16 (4.3) | 9 (2.3) | 10 (1.2) | 163 (3.6) |
| Multiple Ethnicities | 95 (3.3) | 16 (4.3) | 23 (6.0) | 54 (6.5) | 188 (4.2) |
| **Education level** |  |  |  |  |  |
| Less than a high school diploma | 55 (1.9) | 0 (0) | 3 (0.7) | 77 (9.3) | 135 (3.0) |
| High school diploma or equivalent | 365 (12.7) | 11 (2.9) | 12 (3.1) | 160 (19.3) | 548 (12.3) |
| Some post-secondary education but no degree | 447 (15.5) | 50 (13.4) | 39 (10.2) | 179 (21.6) | 715 (16.0) |
| Associates degree | 192 (6.6) | 27 (7.2) | 27 (7.0) | 47 (5.6) | 293 (6.5) |
| Bachelor degree | 853 (29.7) | 120 (32.2) | 149 (39.1) | 144 (17.4) | 1266 (28.4) |
| Graduate degree | 389 (13.5) | 19 (5.1) | 97 (25.4) | 53 (6.4) | 558 (12.5) |
| Registered apprenticeship or other trades<br>certificate or diploma | 463 (16.1) | 143 (38.4) | 51 (13.3) | 156 (18.8) | 813 (18.2) |
| No Education data | 102 (3.5) | 2 (0.5) | 3 (0.7) | 10 (1.2) | 117 (2.6) |
| **Married or Living with Partner** |  |  |  |  |  |
| Has partner | 1672 (58.3) | 256 (68.8) | 204 (53.5) | 371 (44.9) | 2503 (56.3) |
| **Employment** |  |  |  |  |  |
| Full time, 40 or more hours per week | 995 (34.7) | 319 (85.7) | 191 (50.1) | 308 (37.2) | 1813 (40.7) |
| Part time, less than 39 hours per week | 651 (22.7) | 31 (8.3) | 147 (38.5) | 206 (24.9) | 1035 (23.2) |
| Unemployed and looking for work | 147 (5.1) | 2 (0.5) | 11 (2.8) | 109 (13.1) | 269 (6.0) |
| Unable to work | 101 (3.5) | 2 (0.5) | 3 (0.7) | 85 (10.2) | 191 (4.2) |
| Unemployed and NOT looking for work | 151 (5.2) | 4 (1.0) | 7 (1.8) | 40 (4.8) | 202 (4.5) |
| Retired | 709 (24.7) | 12 (3.2) | 19 (4.9) | 67 (8.1) | 807 (18.1) |
| Missing | 112 (3.9) | 2 (0.5) | 3 (0.7) | 11 (1.3) | 128 (2.8) |
| **Total family income** |  |  |  |  |  |
| Less than \$20,000 | 149 (5.1) | 2 (0.5) | 7 (1.8) | 150 (18.1) | 308 (6.9) |
| \$20,000 to \$34,999 | 238 (8.3) | 3 (0.8) | 30 (7.8) | 134 (16.2) | 405 (9.1) |
| \$35,000 to \$49,999 | 302 (10.5) | 11 (2.9) | 30 (7.8) | 112 (13.5) | 455 (10.2) |
| \$50,000 to \$74,999 | 465 (16.2) | 24 (6.4) | 65 (17.0) | 147 (17.7) | 701 (15.7) |
| \$75,000 to \$99,999 | 478 (16.6) | 51 (13.7) | 57 (14.9) | 105 (12.7) | 691 (15.5) |
| \$100,000 to \$149,999 | 517 (18.0) | 123 (33.0) | 90 (23.6) | 94 (11.3) | 824 (18.5) |
| \$150,000 or more | 363 (12.6) | 135 (36.2) | 73 (19.1) | 45 (5.4) | 616 (13.8) |
| Missing | 354 (12.3) | 23 (6.1) | 29 (7.6) | 39 (4.7) | 445 (10.0) |
| **Housing** |  |  |  |  |  |
| I live in a house that I own | 1733 (60.4) | 293 (78.7) | 214 (56.1) | 300 (36.3) | 2540 (57.1) |
| Renting apartment or house | 794 (27.7) | 61 (16.3) | 123 (32.2) | 381 (46.1) | 1359 (30.5) |
| living with a friend or family member | 235 (8.1) | 13 (3.4) | 40 (10.4) | 81 (9.8) | 369 (8.3) |
| Living in supportive housing | 6 (0.2) | 0 (0) | 2 (0.5) | 25 (3.0) | 33 (0.7) |
| Living in a shelter facility | 4 (0.1) | 0 (0) | 0 (0) | 15 (1.8) | 19 (0.4) |
| Currently unhoused | 1 (0.0) | 0 (0) | 0 (0) | 20 (2.4) | 21 (0.4) |
| Missing | 93 (3.2) | 5 (1.3) | 2 (0.5) | 4 (0.4) | 104 (2.3) |
| **Province** |  |  |  |  |  |
| Ontario | 1048 (36.5) | 110 (29.5) | 112 (29.3) | 287 (34.7) | 1557 (35.0) |
| Alberta | 392 (13.6) | 178 (47.8) | 95 (24.9) | 166 (20.0) | 831 (18.6) |
| British Columbia | 358 (12.4) | 22 (5.9) | 44 (11.5) | 96 (11.6) | 520 (11.6) |
| Québec | 276 (9.6) | 20 (5.3) | 31 (8.1) | 74 (8.9) | 401 (9.0) |
| Saskatchewan | 164 (5.7) | 11 (2.9) | 27 (7.0) | 67 (8.1) | 269 (6.0) |
| Manitoba | 160 (5.5) | 2 (0.5) | 18 (4.7) | 38 (4.6) | 218 (4.9) |
| New Brunswick | 164 (5.7) | 3 (0.8) | 23 (6.0) | 28 (3.3) | 218 (4.9) |
| Nova Scotia | 108 (3.7) | 19 (5.1) | 11 (2.8) | 36 (4.3) | 174 (3.9) |
| Newfoundland and Labrador | 104 (3.6) | 3 (0.8) | 12 (3.1) | 14 (1.6) | 133 (2.9) |
| Prince Edward Island | 55 (1.9) | 2 (0.5) | 5 (1.3) | 7 (0.8) | 69 (1.5) |
| Yukon | 25 (0.8) | 0 (0) | 1 (0.2) | 5 (0.6) | 31 (0.6) |
| Northwest Territories | 8 (0.2) | 1 (0.2) | 0 (0) | 3 (0.3) | 12 (0.2) |
| Nunavut | 3 (0.1) | 0 (0) | 0 (0) | 3 (0.3) | 6 (0.1) |
| Missing | 1 (0.0) | 1 (0.2) | 2 (0.5) | 2 (0.2) | 6 (0.1) |
| **Community Size** |  |  |  |  |  |
| Large urban (>100,000 people) | 1452 (50.6) | 193 (51.8) | 198 (51.9) | 455 (55.0) | 2298 (51.6) |
| Medium urban (30,000 - 99,999 people) | 534 (18.6) | 61 (16.3) | 76 (19.9) | 157 (19.0) | 828 (18.6) |
| Small urban (1,000 - 29,999 people) | 456 (15.9) | 76 (20.4) | 70 (18.3) | 119 (14.4) | 721 (16.2) |
| Rural (<1,000 people) | 310 (10.8) | 41 (11.0) | 29 (7.6) | 65 (7.8) | 445 (10.0) |
| Missing | 114 (3.9) | 1 (0.2) | 8 (2.0) | 30 (3.6) | 153 (3.4) |

**Table 4:** Demographics of participants in the test-retest dataset. Unless otherwise specified, numbers are counts with percentage of total respondents in brackets.

| **Key Interest Groups** |  |
| --- | --- |
| PWUS | 12 (8.5) |
| Emergency responders | 1 (0.7) |
| Healthcare providers | 12 (8.5) |
| General public | 117 (82.4) |
| **Age** |  |
| Mean | 48.7 (0) |
| Standard deviation | 17.3 (0) |
| **Gender** |  |
| Male | 61 (43.6) |
| Female | 79 (56.4) |
| Transgender Male | 0 (0) |
| Transgender Female | 0 (0) |
| Gender-diverse | 0 (0) |
| · Non-binary | 0 (0) |
| · Gender Fluid/Queer | 0 (0) |
| · Two-Spirit | 0 (0) |
| No Gender | 0 (0) |
| **Ethnicity** |  |
| White | 126 (83.4) |
| Indigenous | 1 (0.7) |
| Other Ethnicities | 18 (11.9) |
| Multiple Ethnicities | 5 (3.3) |
| Missing | 1 (0.7) |
| **Education level** |  |
| Less than a high school diploma | 1 (0.7) |
| High school diploma or equivalent | 14 (10) |
| Some post-secondary education but no degree | 19 (13.6) |
| Associates degree | 7 (5) |
| Bachelor degree | 45 (32.1) |
| Graduate degree | 26 (18.6) |
| Registered apprenticeship or other trades<br>certificate or diploma | 28 (20) |
| No Education data | 0 (0) |
| **Employment** |  |
| Full time, 40 or more hours per week | 52 (37.1) |
| Part time, less than 39 hours per week | 39 (27.9) |
| Unemployed and looking for work | 6 (4.3) |
| Unable to work | 1 (0.7) |
| Unemployed and NOT looking for work | 4 (2.9) |
| Retired | 37 (26.4) |
| Missing | 1 (0.7) |
| **Total family income** |  |
| Less than \$20,000 | 5 (3.6) |
| \$20,000 to \$34,999 | 13 (9.3) |
| \$35,000 to \$49,999 | 10 (7.1) |
| \$50,000 to \$74,999 | 32 (22.9) |
| \$75,000 to \$99,999 | 19 (13.6) |
| \$100,000 to \$149,999 | 31 (22.1) |
| \$150,000 or more | 20 (14.3) |
| Missing | 10 (7.1) |
| **Housing** |  |
| I live in a house that I own | 84 (60) |
| Renting apartment or house | 47 (33.6) |
| living with a friend or family member | 8 (5.7) |
| Living in supportive housing | 0 (0) |
| Living in a shelter facility | 0 (0) |
| Currently unhoused | 0 (0) |
| Missing | 1 (0.7) |
| **Province** |  |
| Ontario | 47 (33.6) |
| Alberta | 9 (6.4) |
| British Columbia | 16 (11.4) |
| Québec | 27 (19.3) |
| Saskatchewan | 8 (5.7) |
| Manitoba | 8 (5.7) |
| New Brunswick | 4 (2.9) |
| Nova Scotia | 6 (4.3) |
| Newfoundland and Labrador | 8 (5.7) |
| Prince Edward Island | 5 (3.6) |
| Yukon | 1 (0.7) |
| Northwest Territories | 0 (0) |
| Nunavut | 1 (0.7) |
| Missing | 0 (0) |
| **Community Size** |  |
| Large urban (>100,000 people) | 71 (50.7) |
| Medium urban (30,000 - 99,999 people) | 26 (18.6) |
| Small urban (1,000 - 29,999 people) | 23 (16.4) |
| Rural (<1,000 people) | 17 (12.1) |
| Missing | 3 (2.1) |

### Respondents

Respondents were an average age of 45.5 years of age with an approximately equal distribution of men and women. There were also 85 gender-diverse participants. The majority of participants (3,286) identified as white, followed by other ethnicity (669), Indigenous (262), and multiple ethnicity (188). There was a wide distribution of education levels, with PWUS and the general public usually having lower levels of education than first responders and healthcare workers. Most respondents also had a partner and were employed (full or part-time). There was a wide distribution of income levels, though most respondents owned or rented their home, while a few were unhoused, living at a shelter, or residing in supportive housing. Respondents were classified according to Statistics Canada’s urban-rural continuum (35). Most respondants were from large urban centers (2,298), but there were also respondents from medium urban (828), small urban (721), and rural locations (445). Overall response rate was 24% among the general public (response rate could not be calculated for the other informant groups due to the purposive sampling approach).

### Test-Retest Reliability

The results of the test-retest reliability analysis undertaken with a sample from the general public are presented in Table 5 (for paired questions) and Table 6 (for unpaired questions). Paired questions are those questions that were asked twice to participants with the same wording - they were asked once for SCS and once for ORHAs. For example, “How do you think SCS affect crime in a community?” and “How do you think ORHAs affect crime in a community?” There was substantial variability in test-retest reliability across questions. Spearman rank coefficients ranged from 0.08 to 0.66 suggesting negligible to moderate stability in question responses between the baseline and follow-up questionnaires. Kappa coefficients range from 0.15 to 0.58 suggesting slight to substantial agreement between question response between baseline and follow-up. Several questions yielded statistically significant results (p<.05) from Wilcoxon Rank Sum Tests, indicating that responses to these questions differed between the baseline and follow-up questionnaires. Test-retest response rate was 13%.

**Table 5:**
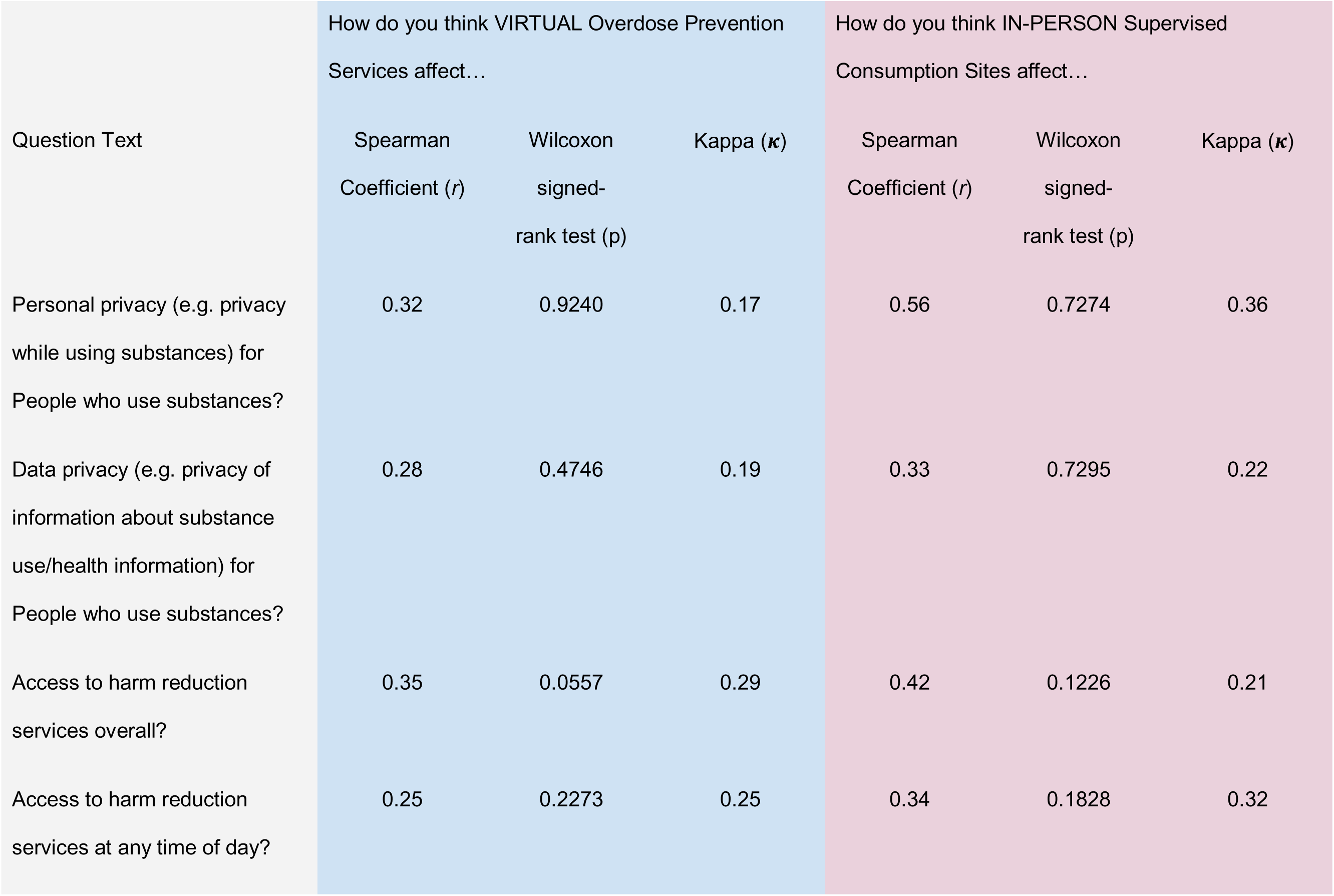
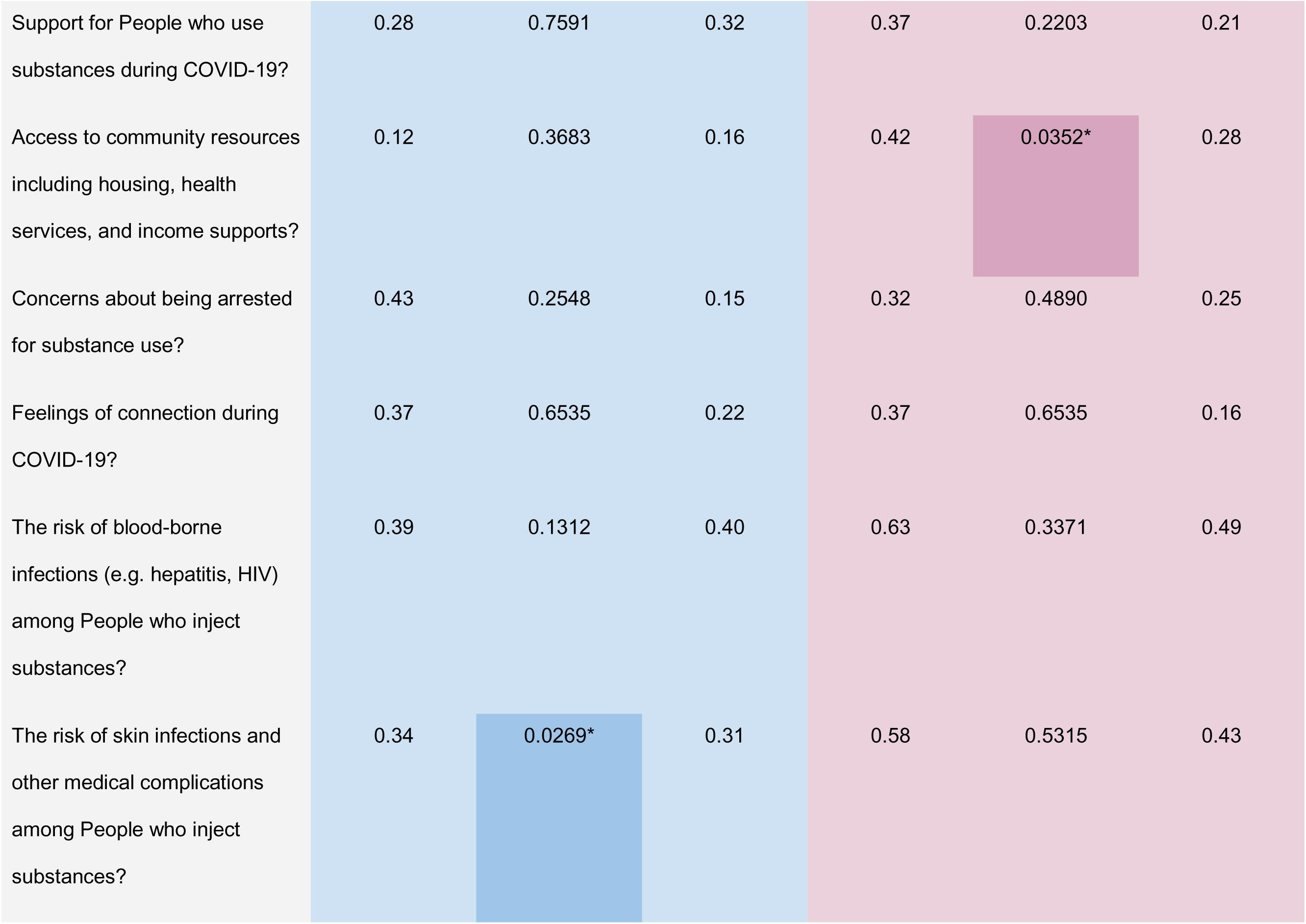
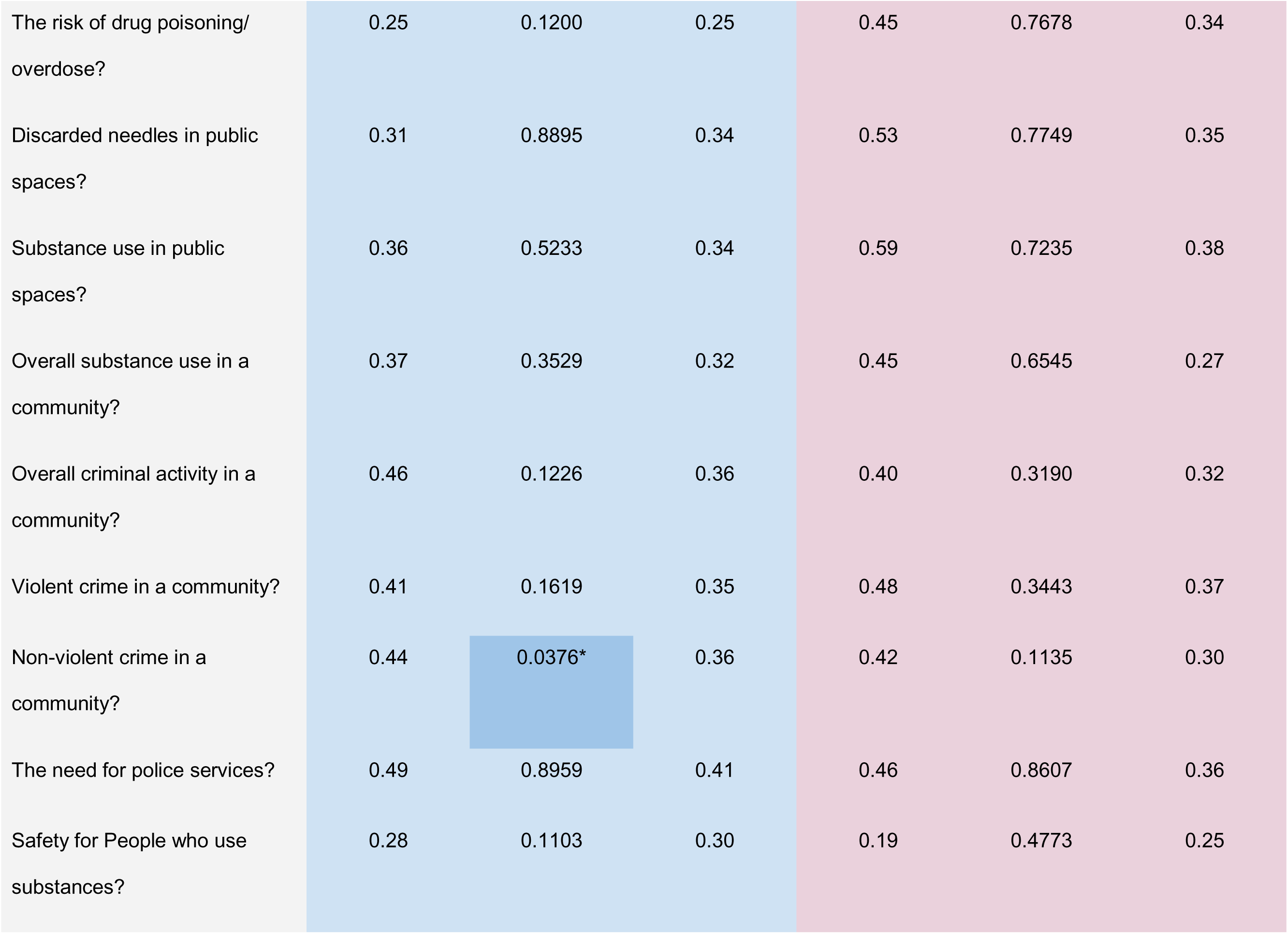
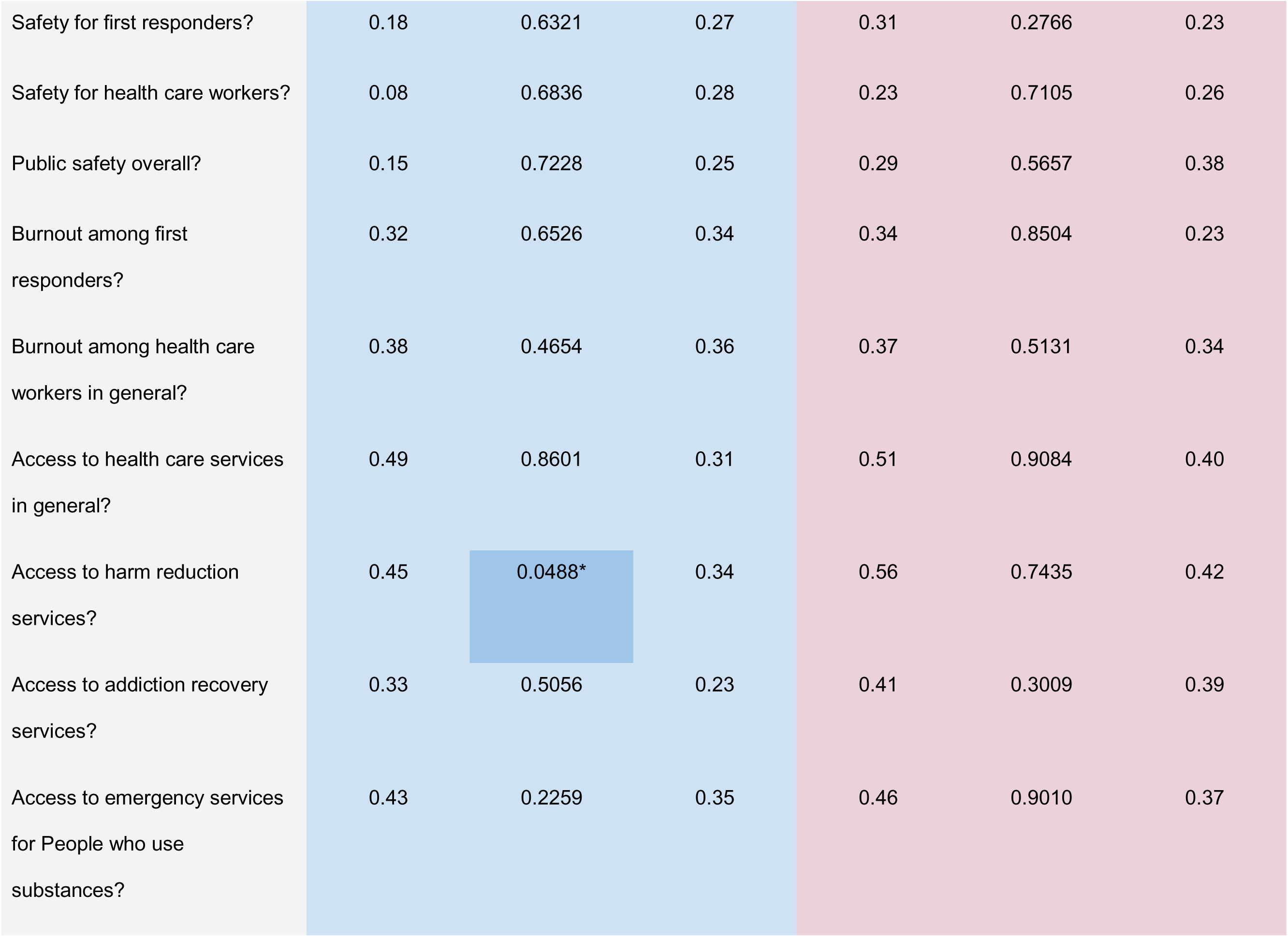
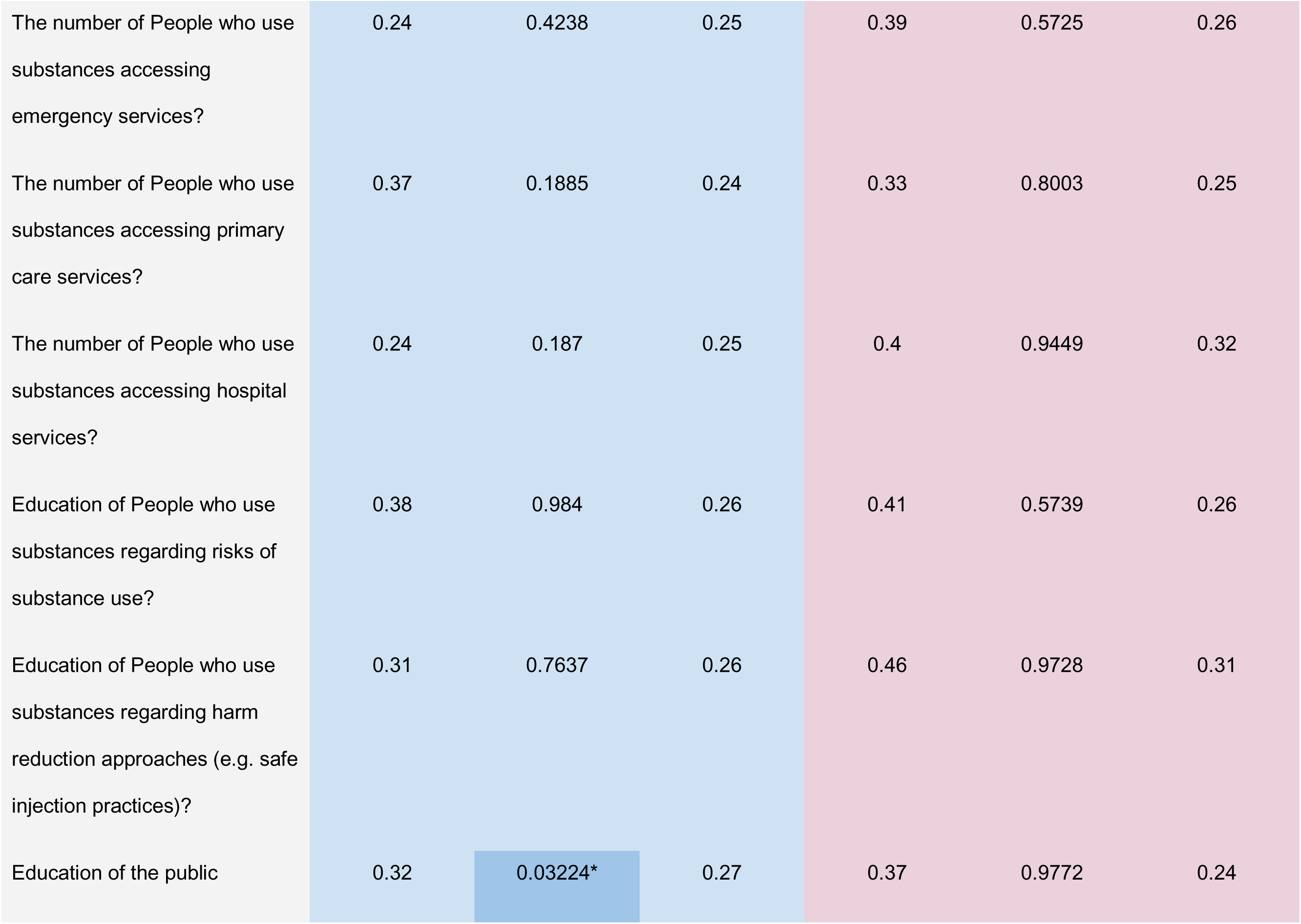
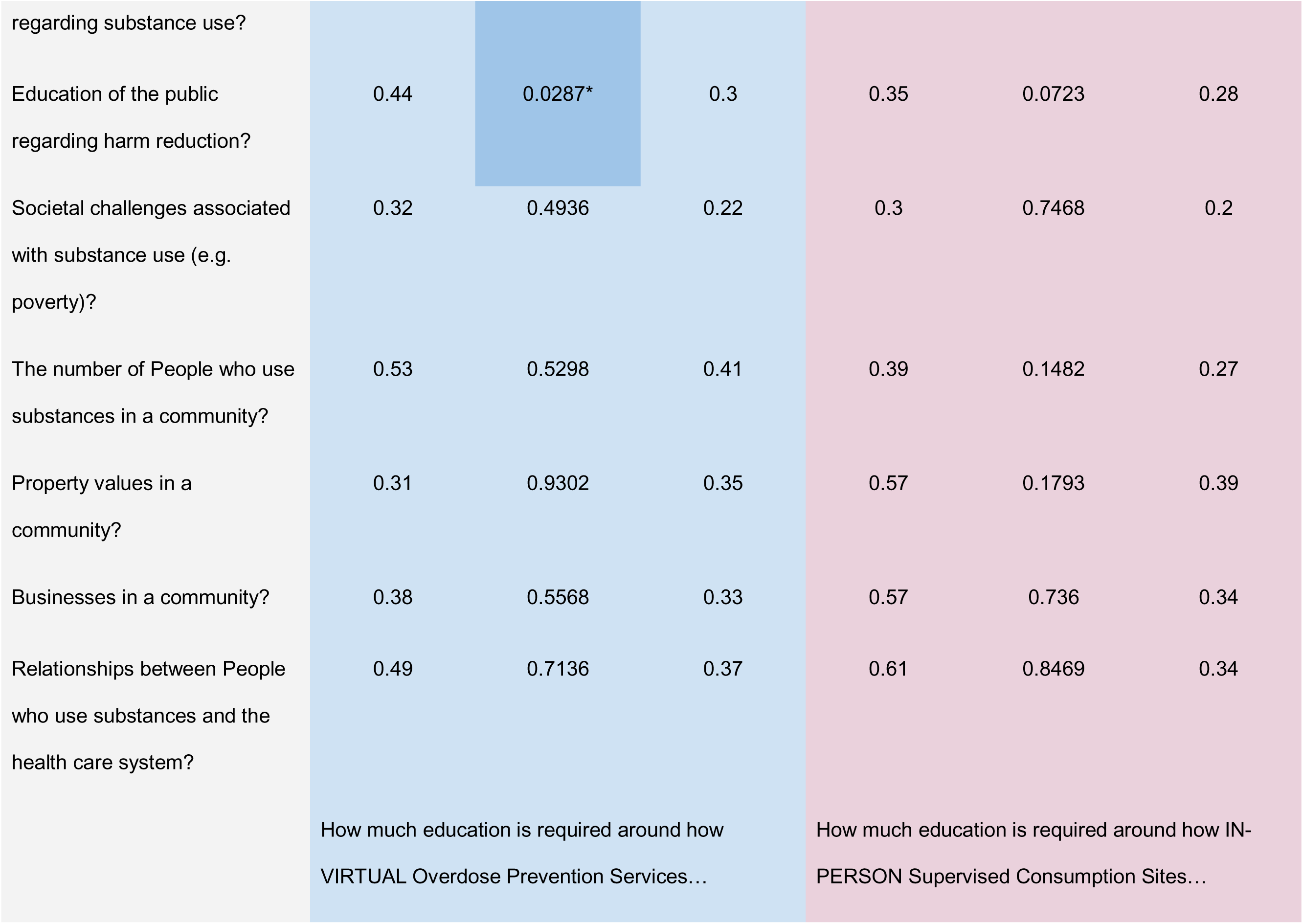
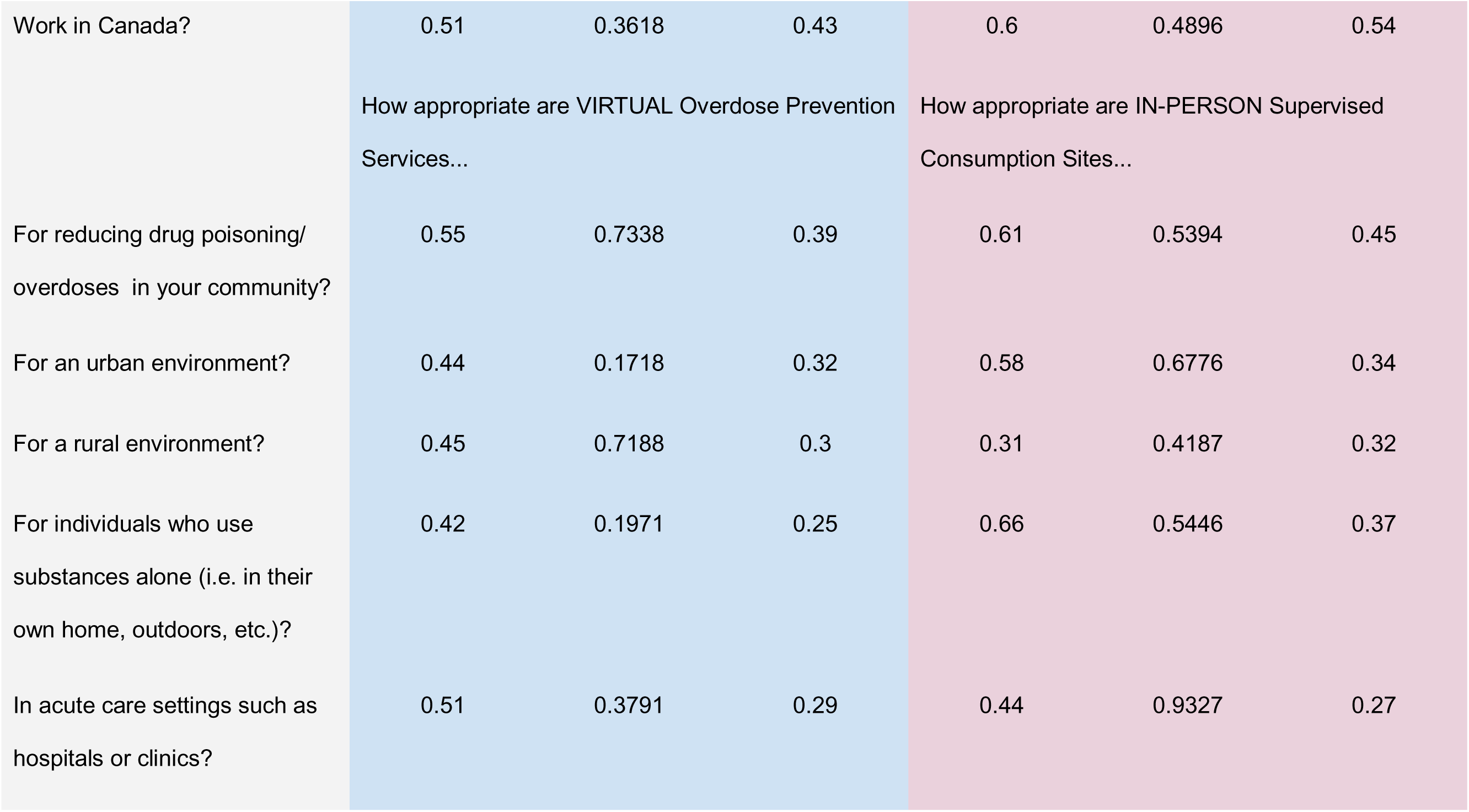
Test-retest reliability analysis for paired questions with a Likert scale (n = 140). Dark shading and asterisks indicate significant results.

**Table 6:** Test-retest reliability analysis for unpaired questions (questions which were not asked about both SCS and ORHAs; n = 140)

|  |  |  |  |
| --- | --- | --- | --- |
| Question Text | Which of the following overdose prevention services would you like to see available or continue to have available in your community? Choose all that apply. |  |  |
| | Spearman Coefficient (r) | Wilcoxon signed-rank test (p) | Kappa ($\kappa$) |
| I want Physical SCS in my community | 0.58 | 0.4413 | 0.58 |
| I want Virtual hotline phone services in my community | 0.41 | 0.0137* | 0.47 |
| I want Virtual app based services in my community | 0.44 | 0.1822 | 0.55 |

## DISCUSSION

The current work is intended to elucidate the questionnaire development, data collection, and reliability analysis components of the CNQOM. This questionnaire is intended to expand on previous work examining attitudes towards SCS while providing a unique perspective on the relationship between attitudes towards SCS and attitudes towards ORHAs. To our knowledge, there has been no other quantitative assessment of key informant attitudes towards ORHAs.

Overall, we were successful in recruiting a diverse group of respondents from the four target informant groups - PWUS, members of the public, healthcare providers, and emergency responders. Due to some difficulty with recruitment of the latter two groups through partner organizations, the general public group was oversampled to help expand the number of respondents in these two groups. Given the potential for online surveys to suffer fraudulent responses, particularly when compensation is offered (36,37), a rigorous response validation procedure was employed, resulting in a robust sample of 4,445 high-quality responses, a number which is similar to or greater than other studies in the field (20,38).

Though our questionnaire is cross-sectional in nature, we decided that it would be in the interest of the reader to conduct a test-retest reliability analysis (39). Our test-retest reliability was lower than what would be considered ideal overall (33,34,40). In particular, seven questions yielded significant Wilcoxon rank sum results, indicating that participant responses were significantly different between measures one and two. In part, the number of significant Wilcoxon results might be explained by the sheer number of comparisons done (approximately 100), of which about five would be expected to be significant by chance. In fact, all significant Wilcoxon results were minimally so (no p values <0.01). During the questionnaire design phase, a number of steps were taken to ensure the face, content, and construct validity of the questions in the instrument. Therefore, we were somewhat surprised by the reliability results for some of the questions. Nonetheless, a number of potential explanations for our relatively low reliability exist.

The CNQOM examines attitudes towards SCS, an intervention which in some cases appears to be poorly understood by policymakers and the public alike (41), and ORHAs, which are a very recent and novel technology in the harm reduction arena (8). Indeed, it is probable that many respondents, especially the general public which largely comprised the test-retest sample, had not heard of ORHAs until they completed our questionnaire. Though we offered participants a brief description of both SCS and ORHAs to ensure a baseline level of knowledge about these interventions, we did not test that knowledge explicitly, and participants did not have much time to think about the interventions when completing the CNQOM. Test-retest reliability is well-known to depend on question content and psychological properties (42). In particular, familiarity and the personal nature of the material have a significant impact on test-retest reliability measures, with greater familiarity and personalness being associated with higher reliability (42). Thus, participant attitudes towards these services might not be particularly stable or their perspectives could evolve, even during the week between our two test-retest reliability measures. Since members of the general public are probably the least informed about ORHAs and SCS (compared to PWUS, first responders, and healthcare providers), it stands to reason that their opinions might be less stable than those of the other groups. Consequently, we hypothesize that test-retest reliability would be higher for groups more familiar with the studied services. Should funding allow, future work might examine the test-retest reliability of the CNQOM among other key informant groups, particularly if the questionnaire is used to assess changes in key informant attitudes over time in the future.

### Overall Strengths

The CNQOM represents the first quantitative examination of key informant attitudes towards ORHAs and one of relatively few quantitative examinations of key informant attitudes towards SCS on a national level. Our large sample size, broad geographic catchment area, diversity of recruitment methods, and inclusion of four key informant groups are major strengths of this study.

### Overall Limitations

A number of the limitations of the CNQOM pertain to the online nature of its administration. Firstly, response bias is an issue with online surveys (43), particularly when compensation is involved (only peers and the general public received compensation, and the amount differed between these groups). Our use of a research analytics company might have reduced response bias among the general public. Secondly, our use of a digital format likely excluded participants who lacked access to electronic devices and an internet connection. This may have caused some degree of sample bias, particularly among PWUS, some of whom may lack access to these technologies (44,45). Thirdly, due to our purposive sampling method using partner organizations, it was not possible to accurately calculate an overall response rate for the questionnaire. Fourthly, it was not possible to conduct test-retest reliability with each stakeholder group due to funding constraints. Nonetheless, we believe that test-retest reliability would likely be better among the other groups than among the general public.

## CONCLUSIONS AND FUTURE DIRECTIONS

This paper documents the methodology of the CNQOM from inception to reliability assessment. The CNQOM quantitatively compares attitudes towards and between SCSs and ORHAs. The questionnaire documents the opinions of four key informant groups - PWUS, the general public, first responders, and healthcare providers. Additionally, it contains novel assessments of ORHA barriers to implementation, methods of promotion, key features, and more. A test-retest reliability analysis demonstrated overall fair to moderate reliability consistent with the content familiarity and personalness of the questions to the test-retest reliability sample of the general public. Data from this questionnaire will help to inform ORHA policy by providing a unique multi-key informant perspective on these novel technologies.

Further work will examine the data from the CNQOM in more detail, including identifying differences in attitudes between SCS and ORHAs, barriers to ORHA use, ways to promote ORHAs, key features of ORHAs, and more. Should the questions be grouped by potential underlying constructs in the future (e.g. safety, community effects, privacy), an analysis of internal consistency or factorial validity should be undertaken (Trakman et al., 2017). The results of this questionnaire are expected to inform quality improvement among existing ORHAs, as well as help guide policy and decision makers on the value of ORHAs.

## Data Availability

Data cannot be shared publicly because of University of Calgary ethics rules. Data are available from the University of Calgary for researchers who meet the criteria for access to confidential data.

## ACKNOWLEDGEMENTS

We would like to acknowledge the financial support provided by Health Canada’s Substance Use and Addictions Program (SUAP) and the Canadian Institutes of Health Research (CIHR) for this research. The Alberta Strategy for Patient Oriented Research Support Unit (AbSPORU) performed data analysis and database creation. Special thanks to Lisa Morris-Miller and Pamela E. Taplay for their subject matter expertise, assistance with validation, and feedback during pilot testing. We also appreciate the assistance of the following entities in participant recruitment: National Overdose Response Service (NORS), Grenfell Ministries, City of Burnaby Police, Kentville Police Service, Kawartha Lakes Police Service, Lower Mainland District RCMP, Calgary Police Service, Medicine Hat Police Service, Brandon Police Service, Edmonton Police Service, Strathroy-Caradoc Police Service, Central Saanich Police Service, West Grey Police Service, Alberta RCMP, Urgences-santé, Northwest Firefighters Association, Government of Yukon Protective Services, New Brunswick Association of Fire Chiefs, City of Yellowknife, Welland Fire and Emergency Services, Calgary Fire Department, Ontario Paramedic Association, Cochrane District EMS, Yukon Government EMS, Halton Health Region Paramedic Services, Saskatchewan College of Paramedics, EHS Nova Scotia, College of Paramedics of Nova Scotia, Alberta Health Services EMS, Island Health, Doctors of Nova Scotia, Saskatchewan Medical Association, Thompson Rivers University Faculty of Social Work, College of Physiotherapists of Ontario, Safeworks, ARCH, MIELS-QUÉBEC, and CIPTO. Their contributions were instrumental in the success of this project.

## REFERENCES

1. Snowdon J, Choi N. Unanticipated Changes in Drug Overdose Death Rates in Canada During the Opioid Crisis. Int J Ment Health Addict [Internet]. 2022 Oct 10 [cited 2023 Sep 12]; Available from: 10.1007/s11469-022-00932-9

2. Belzak L, Halverson J. The opioid crisis in Canada: a national perspective. Health Promot Chronic Dis Prev Can Res Policy Pract. 2018 Jun;38(6):224–33.

3. Mattson CL, O’Donnell J, Kariisa M, Seth P, Scholl L, Gladden RM. Opportunities to Prevent Overdose Deaths Involving Prescription and Illicit Opioids, 11 States, July 2016-June 2017. MMWR Morb Mortal Wkly Rep. 2018 Aug 31;67(34):945–51.

4. Gicquelais RE, Genberg BL, Maksut JL, Bohnert ASB, Fernandez AC. Prevalence and correlates of using opioids alone among individuals in a residential treatment program in Michigan: implications for overdose mortality prevention. Harm Reduct J. 2022 Dec 3;19(1):135.

5. Norton A, Hayashi K, Johnson C, Choi J, Milloy MJ, Kerr T. Injecting drugs alone during an overdose crisis in Vancouver, Canada. Harm Reduct J. 2022 Nov 17;19(1):125.

6. Foreman-Mackey A, Xavier J, Corser J, Fleury M, Lock K, Mehta A, et al. “It’s just a perfect storm”: Exploring the consequences of the COVID-19 pandemic on overdose risk in British Columbia from the perspectives of people who use substances. BMC Public Health. 2023 Apr 3;23(1):640.

7. Matskiv G, Marshall T, Krieg O, Viste D, Ghosh SM. Virtual overdose monitoring services: a novel adjunctive harm reduction approach for addressing the overdose crisis. CMAJ Can Med Assoc J J Assoc Medicale Can. 2022 Nov 28;194(46):E1568–72.

8. Loverock A, Marshall T, Viste D, Safi F, Rioux W, Sedaghat N, et al. Electronic harm reduction interventions for drug overdose monitoring and prevention: A scoping review. Drug Alcohol Depend. 2023 Sep 1;250:110878.

9. Proctor E, Silmere H, Raghavan R, Hovmand P, Aarons G, Bunger A, et al. Outcomes for Implementation Research: Conceptual Distinctions, Measurement Challenges, and Research Agenda. Adm Policy Ment Health. 2011;38(2):65–76.

10. Marshall T, Viste D, Jones S, Kim J, Lee A, Jafri F, et al. Beliefs, attitudes and experiences of virtual overdose monitoring services from the perspectives of people who use substances in Canada: a qualitative study. Harm Reduct J. 2023 Jun 24;20(1):80.

11. Tas B, Lawn W, Traykova EV, Evans RAS, Murvai B, Walker H, et al. A scoping review of mHealth technologies for opioid overdose prevention, detection and response. Drug Alcohol Rev. 2023;42(4):748–64.

12. Rider N, Safi F, Marshall T, Jones S, Seo B, Viste D, et al. Investigating uses of peer-operated Virtual Overdose Monitoring Services (VOMS) beyond overdose response: a qualitative study. Am J Drug Alcohol Abuse. 2023 Nov 2;49(6):809–17.

13. Rioux W, Marshall T, Ghosh SM. Virtual overdose monitoring services and overdose prevention technologies: Opportunities, limitations, and future directions. Int J Drug Policy. 2023 Sep;119:104121.

14. Seo B, Rider N, Rioux W, Teare A, Jones S, Taplay P, et al. Understanding the barriers and facilitators to implementing and sustaining Mobile Overdose Response Services from the perspective of Canadian key interest groups: a qualitative study. Harm Reduct J. 2024 Feb 2;21(1):28.

15. Lange BCL, Bach-Mortensen AM. A systematic review of stakeholder perceptions of supervised injection facilities. Drug Alcohol Depend. 2019 Apr 1;197:299–314.

16. Kennedy MC, Karamouzian M, Kerr T. Public Health and Public Order Outcomes Associated with Supervised Drug Consumption Facilities: a Systematic Review. Curr HIV/AIDS Rep. 2017 Oct;14(5):161–83.

17. Kerr T, Mitra S, Kennedy MC, McNeil R. Supervised injection facilities in Canada: past, present, and future. Harm Reduct J. 2017 May 18;14(1):28.

18. Levengood TW, Yoon GH, Davoust MJ, Ogden SN, Marshall BDL, Cahill SR, et al. Supervised Injection Facilities as Harm Reduction: A Systematic Review. Am J Prev Med. 2021 Nov;61(5):738–49.

19. Potier C, Laprévote V, Dubois-Arber F, Cottencin O, Rolland B. Supervised injection services: what has been demonstrated? A systematic literature review. Drug Alcohol Depend. 2014 Dec 1;145:48–68.

20. Wild TC, Koziel J, Anderson-Baron J, Asbridge M, Belle-Isle L, Dell C, et al. Public support for harm reduction: A population survey of Canadian adults. PloS One. 2021;16(5):e0251860.

21. Eykelenboom M, van Stralen MM, Olthof MR, Schoonmade LJ, Steenhuis IHM, Renders CM, et al. Political and public acceptability of a sugar-sweetened beverages tax: a mixed-method systematic review and meta-analysis. Int J Behav Nutr Phys Act. 2019 Sep 4;16(1):78.

22. Galatas B, Nhantumbo H, Soares R, Djive H, Murato I, Simone W, et al. Community acceptability to antimalarial mass drug administrations in Magude district, Southern Mozambique: A mixed methods study. PLoS ONE. 2021 Mar 23;16(3):e0249080.

23. Ridgeway K, Montgomery ET, Smith K, Torjesen K, van der Straten A, Achilles SL, et al. Vaginal ring acceptability: A systematic review and meta-analysis of vaginal ring experiences from around the world. Contraception. 2022 Feb;106:16–33.

24. Scheidmeir M, Kubiak T, Luszczynska A, Wendt J, Scheller DA, Meshkovska B, et al. Acceptability of policies targeting dietary behaviours and physical activity: a systematic review of tools and outcomes. Eur J Public Health. 2022 Nov 29;32(Suppl 4):iv32–49.

25. Trakman GL, Forsyth A, Hoye R, Belski R. Developing and validating a nutrition knowledge questionnaire: key methods and considerations. Public Health Nutr. 2017 Oct;20(15):2670– 9.

26. Clark LA, Watson D. Constructing validity: New developments in creating objective measuring instruments. Psychol Assess. 2019 Dec;31(12):1412–27.

27. Story DA, Tait AR. Survey Research. Anesthesiology. 2019 Feb;130(2):192–202.

28. Statistics Canada. The Daily — Canada’s population estimates, third quarter 2023 [Internet]. 2023 [cited 2024 Nov 28]. Available from: https://www150.statcan.gc.ca/n1/daily-quotidien/231219/dq231219c-eng.htm

29. Government of Canada. Canadian Occupational Projection System [Internet]. [cited 2024 Nov 28]. Available from: https://occupations.esdc.gc.ca/sppc-cops/l.3bd.2t.1ils@-eng.jsp?lid=85

30. Health Canada. Canadian Alcohol and Drugs Survey (CADS): summary of results for 2019 [Internet]. 2021 [cited 2024 Nov 28]. Available from: https://www.canada.ca/en/health-canada/services/canadian-alcohol-drugs-survey/2019-summary.html

31. Bennett ME, Nidecker M, Kinnaman JES, Li L, Bellack AS. Examination of the Inventory of Drug Use Consequences with Individuals with Serious and Persistent Mental Illness and Co-occurring Substance Use Disorders. Am J Drug Alcohol Abuse. 2009;35(5):385.

32. Spirito A, Bromberg JR, Casper TC, Chun TH, Mello MJ, Dean JM, et al. Reliability and Validity of a Two-Question Alcohol Screen in the Pediatric Emergency Department. Pediatrics. 2016 Dec;138(6):e20160691.

33. Landis JR, Koch GG. The measurement of observer agreement for categorical data. Biometrics. 1977 Mar;33(1):159–74.

34. Mukaka MM. Statistics corner: A guide to appropriate use of correlation coefficient in medical research. Malawi Med J J Med Assoc Malawi. 2012 Sep;24(3):69–71.

35. Statistics Canada. Illustrated Glossary - Population centre (POPCTR) [Internet]. 2017 [cited 2024 Nov 24]. Available from: https://www150.statcan.gc.ca/n1/pub/92-195-x/2021001/geo/pop/pop-eng.htm

36. Ball HL. Conducting Online Surveys. J Hum Lact Off J Int Lact Consult Assoc. 2019 Aug;35(3):413–7.

37. Palamar JJ, Acosta P. On the efficacy of online drug surveys during the time of COVID-19. Subst Abuse. 2020;41(3):283–5.

38. Kuo M, Shamsian A, Tzemis D, Buxton JA. A drug use survey among clients of harm reduction sites across British Columbia, Canada, 2012. Harm Reduct J. 2014 Apr 27;11(1):13.

39. Rattray J, Jones MC. Essential elements of questionnaire design and development. J Clin Nurs. 2007 Feb;16(2):234–43.

40. McHugh ML. Interrater reliability: the kappa statistic. Biochem Medica. 2012;22(3):276–82.

41. Salvalaggio G, Brooks H, Caine V, Gagnon M, Godley J, Houston S, et al. Flawed reports can harm: the case of supervised consumption services in Alberta. Can J Public Health Rev Can Santé Publique. 2023 Nov 6;114(6):928–33.

42. Brackbill Y. Test-Retest Reliability in Population Research. Stud Fam Plann. 1974;5(8):261– 6.

43. Wright KB. Researching Internet-Based Populations: Advantages and Disadvantages of Online Survey Research, Online Questionnaire Authoring Software Packages, and Web Survey Services. J Comput-Mediat Commun. 2005 Apr 1;10(3):JCMC1034.

44. Milward J, Day E, Wadsworth E, Strang J, Lynskey M. Mobile phone ownership, usage and readiness to use by patients in drug treatment. Drug Alcohol Depend. 2015 Jan 1;146:111– 5.

45. Ozga JE, Paquette C, Syvertsen JL, Pollini RA. Mobile phone and internet use among people who inject drugs: Implications for mobile health interventions. Subst Abuse. 2022;43(1):592–7.

